# Beta-HCG levels and ovarian ultrasonography results among non-pregnant women of reproductive age in Port Harcourt, Nigeria

**DOI:** 10.1101/2023.07.05.23292265

**Authors:** Celine Chibuzo Agonsi, Francis Anacletus, Joel Aluko, Chinemerem Eleke, Joy Samuel

**Affiliations:** African Centre of Public Health and Toxicological Research (ACE-PUTOR), University of Port Harcourt, Choba, Nigeria; Department of Nursing Science, University of Port Harcourt, Choba, Nigeria

**Author notes:** **Corresponding Author** Chinemerem Eleke, (CE). **Authors’ contributions** All authors (CCA, FA, JA, and CE) contributed to the conceptualization, data curation, drafting of the original manuscript, and critical revision of the final manuscript. All authors read and approved the final version of the manuscript.

**Keywords:** Pregnancy, ovarian neoplasm, ultrasonography, biomarkers

## Abstract

**Background:** Certain ovarian cancers previously more common in postmenopausal women are now increasingly observed in women of reproductive age. The research on using β-HCG as a diagnostic biomarker for ovarian cancer in women of reproductive age is ongoing. This particular study assessed the level of serum β-HCG in non-pregnant women of reproductive age and determined its potential association with suspicious ovarian ultrasonography results in Port Harcourt, Nigeria.

**Material and methods:** This study utilized a descriptive-analytic design on a quota sample of 224 diagnostic case notes of women aged 18-40 years obtained from eight diagnostic centres. Data collection involved a data extraction form. Data analysis employed descriptive statistics, Chi-square, Fisher’s exact test, and Odds Ratio at 95% confidence and 5% significance levels.

**Results:** About 5.8% of the participants exhibited detectable levels of serum β-HCG above 5 IU/L (World Health Organization reference) at a mean concentration of 5.87 (±1.75) IU/L. About 4.0% of the participants had suspicious ovarian lesions identified through ultrasonography. Participants with elevated serum β-HCG levels above the WHO reference were 59 times more likely to have suspicious ovarian lesions, with an odds ratio of 59.4 (95%CI: 12.3–287.8, p = 0.001). There was a significant association between serum β-HCG level and age (p = 0.041) as well as parity (p < 0.001).

**Conclusions:** This study demonstrated that Serum β-HCG levels above the WHO reference were associated with suspicious ovarian lesions. Non-pregnant women should undergo serum β-HCG testing at least yearly to facilitate the early detection of ovarian anomalies.

## Introduction

Ovarian cancer is one of the most common cancers and a leading cause of mortality among women [1]. It is often diagnosed at an advanced stage, resulting in poor health outcomes. Previous studies have suggested that elevated serum levels of gonadotropins, including β-HCG, may be associated with ovarian cancer [2-4]. However, the use of β-HCG as a diagnostic biomarker in women of reproductive age remains controversial due to various concerns and limitations [5].

The β-HCG is a glycoprotein known to be synthesized by the foetal trophoblast and released into maternal blood. Nonetheless, recent research report that some cancers do produce β-HCG [2, 6]. Clinical laboratory protocols enable the measurement of β-HCG in blood or serum specimens [7]. Initially used for diagnosing and monitoring pregnancy, some studies support its potential as a biomarker for certain cancers, including ovarian cancer [8, 9].

The concerns surrounding β-HCG as a diagnostic biomarker for ovarian cancer are palpable. Firstly, previous studies have observed elevated β-HCG levels in a small percentage of healthy non-pregnant women of reproductive age without detectable ovarian cancer [10]. Secondly, serum β-HCG levels increase with age and parity status, making it challenging to establish standardized cut off values [11]. Moreover, there is a lack of consensus among laboratories regarding the cut off values, leading to significant variation in reporting quantitative results [7]. Additionally, the reference limits suggested by manufacturers of commercial HCG assay kits may not align with actual clinical practice [12]. Despite these limitations, some studies support the use of β-HCG as a biomarker for ovarian cancer, particularly in advanced stages [8, 9].

Non-pregnant women of reproductive age (specifically those aged 18-40 years) have scarcely been examined by previous studies regarding the prevalence of anomalous ovarian lesions and their association with elevated serum levels of β-HCG. The dearth of literature highlights the need for further research on the subject matter. Understanding the potential of β-HCG as a diagnostic biomarker for ovarian disease in non-pregnant women of reproductive age is crucial for the early detection of ovarian neoplasm. This study aims to investigate the serum β-HCG levels and prevalence of suspicious ovarian lesions among non-pregnant women of reproductive age in Port Harcourt, Nigeria. The findings of this study may contribute to the ongoing discourse surrounding the use of β-HCG as a biomarker for ovarian cancer.

## Material and methods

### Ethics statement

The University of Port Harcourt Institutional Review Board granted approval for this study, with the approval number UPH/CEREMAD/REC/MM87/068. The study followed the guidelines outlined in the Helsinki Declaration of 1975, revised in 2013. Written consent was not obtained as the study reviewed anonymous (de-identified) diagnostic case notes. The study was conducted between November 2022 and May 2023. Recruitment of participant case notes began February 2023.

### Study area and period

This study examined case notes in private diagnostic centres in Port Harcourt, Rivers State, Nigeria. Located in the oil-rich Niger Delta, Port Harcourt is the capital of Rivers State. At least 2.4 million persons are residents in the city and engage in various occupations such as construction, trading, and white and blue-collar jobs. However, Port Harcourt has environmental challenges, including the heavy burning of fossil fuels and the resulting soot pollution from clandestine illegal crude oil processing in neighbouring communities. Port Harcourt has many private diagnostic centres, but only eight demonstrated sufficient technology to test and quantify Human Chorionic Gonadotropin. The centres include De-Integrated Medical Diagnostic and Research Laboratory, Asi Ukpo Medical Imaging and Phlebotomy Centre, Bio-systems Medical Diagnostics, Dorek Medical Diagnostics, Pyramids Diagnostics, Index Medical and Diagnostics, Trans-view Diagnostics, Oasis Diagnostic Centre.

### Study design and population

This study utilized a descriptive-analytic design to investigate a specific research question. The target population for this study consisted of approximately 301,562 women who are residents of Port Harcourt and fall within the age range of 18 to 40 years old.

### Sample size determination and sampling procedure

The sample size for this study, consisting of women of reproductive age, was determined using the Cochran’s formula [13]. The formula, mathematically represented as n = [(Z)2 x P(1-P)÷d2], took into account various factors. These included the critical value constant at a power of 80% (Z = 1.96), the estimated prevalence based on literature (P = 84%) [14], and the tolerable error level (d = 5%). The computed minimum sample size was 206. To address potential non-response, the minimum sample size was increased by 10% using the non-response formula [15], expressed as n* = [n ÷ (1-0.1)]. This resulted in a final sample size of 224 participants. The selection of case notes from the participating diagnostic centres involved the application of a quota sampling technique. A total of 28 diagnostic case notes were chosen from each of the eight centres (n = 224). Women who were aged 18-40 years were included. On the other hand, women who were pregnant and residing outside the Port Harcourt Metropolis area were excluded.

### Instrument and data collection

The research team utilized a semi-structured data extraction form consisting of six items to collect relevant data. The form was divided into three sections, namely A, B, and C. Section A gathered background demographic information of the participants, such as age, marital status, parity status, and employment status, using five instrument items. Section B focused on extracting the laboratory results for serum HCG concentration. Section C extracted the results of ovarian ultrasonography.

Data were extracted from the selected case notes. Demographic data of the participants were analyzed using descriptive statistical methods, including frequency and percentage. The interval data related to age were summarized using measures such as mean, standard deviation, frequency, and percentage. The association between variables was examined using statistical tests such as the Chi-square test, Fisher exact test, and Odds ratio. These inferential statistics were conducted at a 95% confidence level and a significance level of 5%. The Statistical Products and Service Solutions software version 25 (IBM, Chicago, IL, USA) was used for all the statistical analyses.

## Results

Table 1 summarises the demographic characteristics of the participants and shows that many (52.7%) of the participants in the study fell within the age range of 30-35 years. The majority of the participants were married (98.7%), had given birth to two or more children (multiparous, 78.6%), and were employed (83.9%).

**Table 1:**
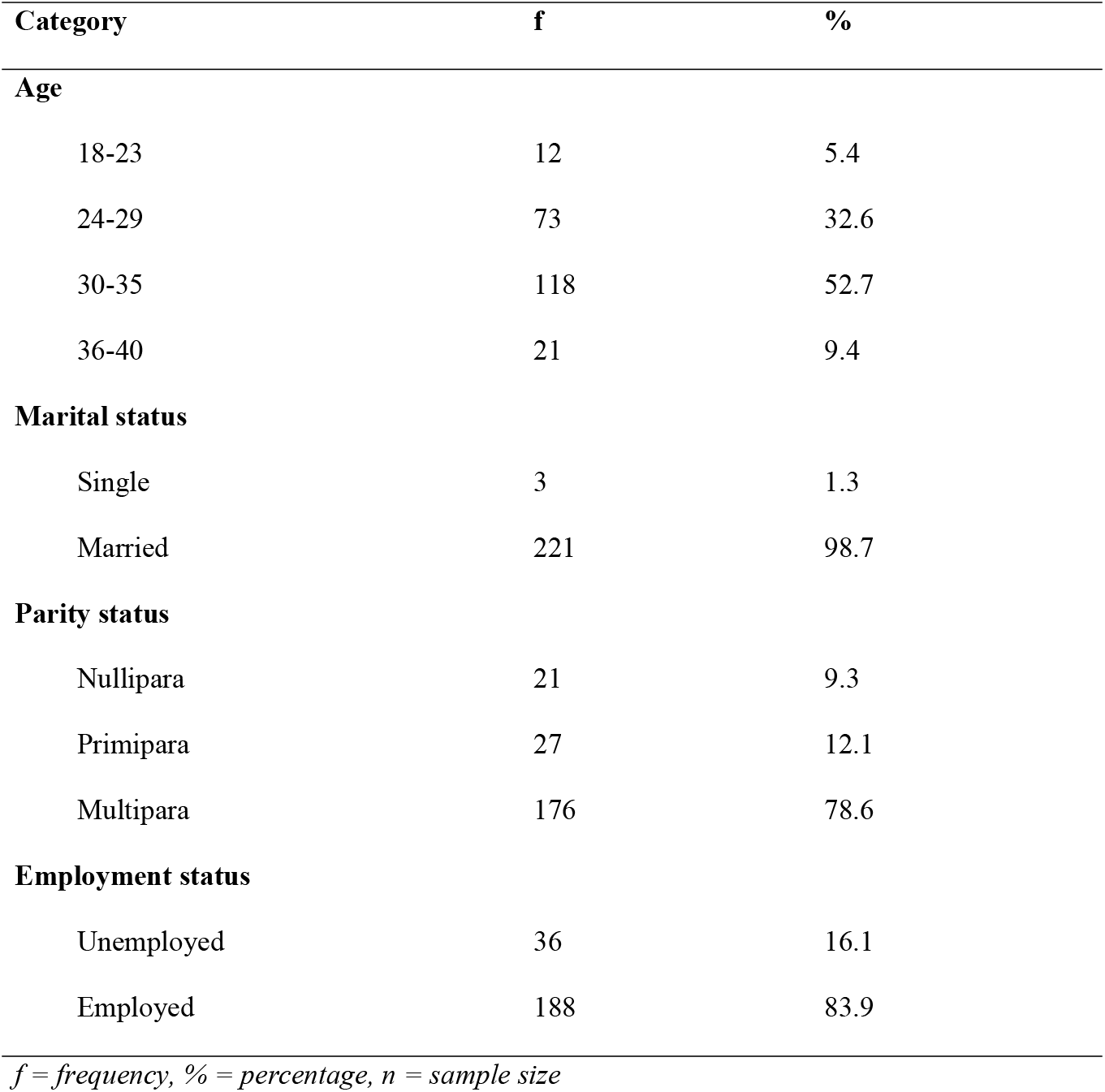
Socio-demographic characteristics of the study participants, (n = 224)

Table 2 summarizes serum β-HCG levels among the participants and indicates that a small proportion (5.8%) of the study participants had a detectable level of serum β-HCG above 5 IU/L, which is the reference value according to the World Health Organization (WHO).

**Table 2:** Serum concentration of serum β-HCG among non-pregnant women, (n = 224)

| Concentration category | f | % | Range | Mean (SD) |
| --- | --- | --- | --- | --- |
| <1.0 IU/L | 4 | 1.8 | - | - |
| 1.0-5.0 IU/L | 207 | 92.4 | 1.1 – 4.6 | 2.95 (0.78) |
| >5.0 IU/L | 13 | 5.8 | 5.3 – 7.1 | 5.87 (1.75) |
| Grand Mean |  |  | 1.1 – 7.1 | 2.96 (1.32) |

*f = frequency, % = percentage, n = sample size, SD = standard deviation*

Figure 1 reveals that few (4.0%) of the study participants were observed to have suspicious ovarian lesions identified during the ultrasonography.

**Figure 1:**
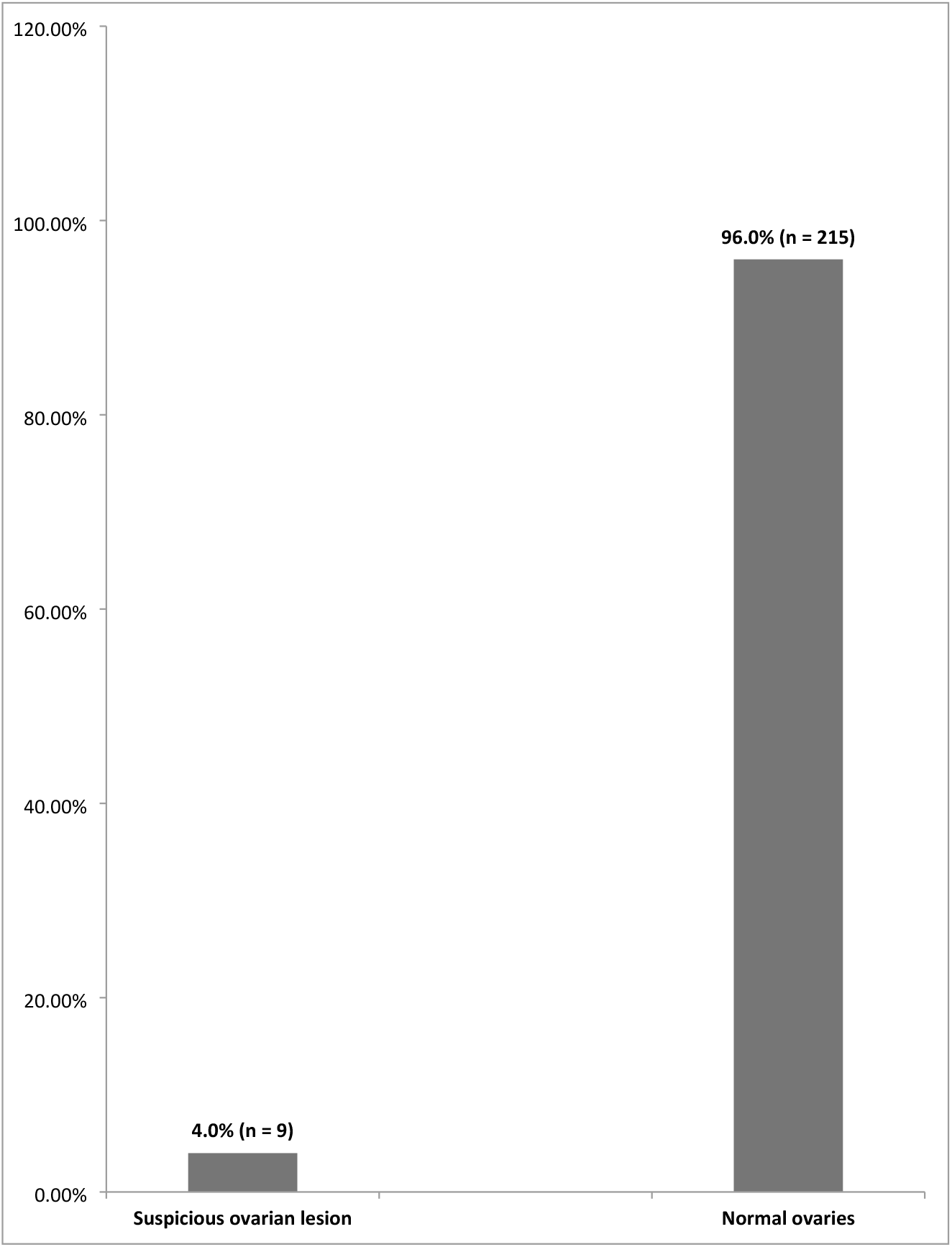
Prevalence of suspicious ovarian lesion among the non-pregnant women

Table 3 summarizes the association between Serum β-HCG concentration and ovarian ultrasonography findings and demonstrates that participants who had elevated serum HCG (> 5IU/L) where 59 folds more likely to have a suspicious ovarian lesion (p = <0.001).

**Table 3:**
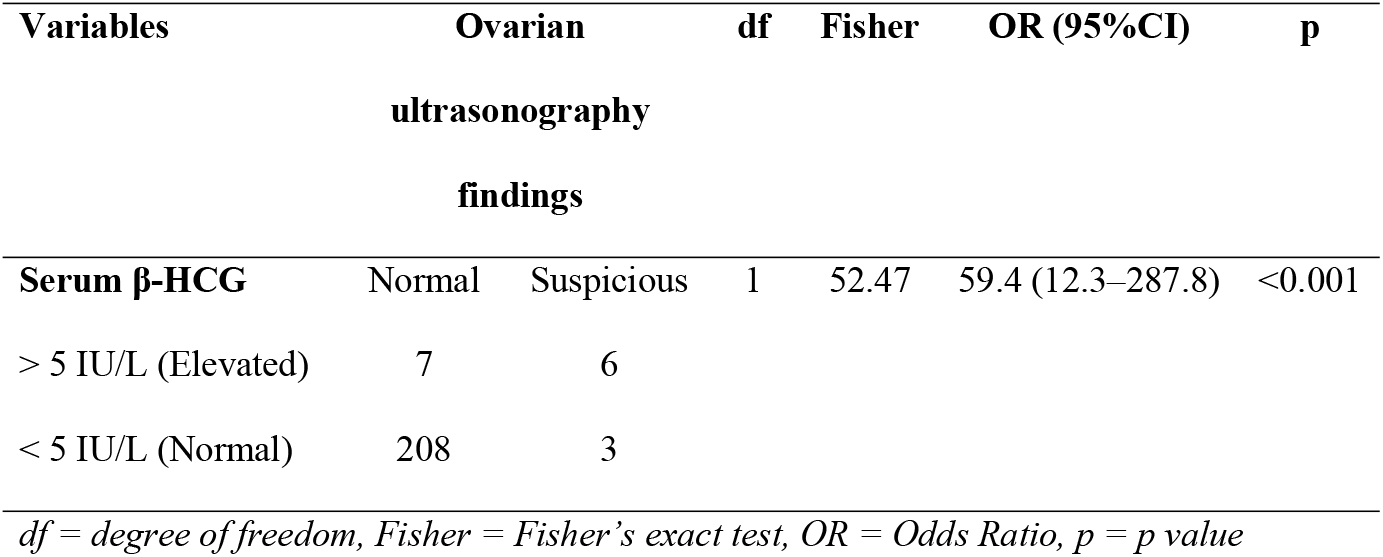
Fisher’s exact test of association between serum HCG and Ovarian ultrasound findings, (n = 224)

| **Variables** | **Ovarian**<br>**ultrasonography**<br>**findings** |  | **df** | **Fisher** | **OR (95%CI)** | **p** |
| --- | --- | --- | --- | --- | --- | --- |
| **Serum β-HCG** | Normal | Suspicious | 1 | 52.47 | 59.4 (12.3–287.8) | <0.001 |
| > 5 IU/L (Elevated) | 7 | 6 |  |  |  |  |
| < 5 IU/L (Normal) | 208 | 3 |  |  |  |  |

*df = degree of freedom, Fisher = Fisher's exact test, OR = Odds Ratio, p = p value*

Table 4 summarizes the association between age and parity with Serum β-HCG concentration and demonstrates a significant association between serum β-HCG level and age (p = 0.041) as well as parity (p < 0.001).

**Table 4:** Chi square test of association between age and parity with HCG concentration, (n = 224)

| **Variables** | **Serum β-HCG concentration** |  | **df** | **$\chi^2$** | **p** |
| --- | --- | --- | --- | --- | --- |
| **Age** | < 5 IU/L (Normal) | > 5 IU/L (Elevated) | 3 | 8.24 | 0.041 |
| 18-23 | 12 | 0 |  |  |  |
| 24-29 | 71 | 2 |  |  |  |
| 30-35 | 111 | 7 |  |  |  |
| 36-40 | 17 | 4 |  |  |  |
| **Parity** |  |  |  |  |  |
| Nullipara | 12 | 9 | 2 | 62.96 | <0.001 |
| Primipara | 24 | 3 |  |  |  |
| Multipara | 175 | 1 |  |  |  |

*df = degree of freedom, $\chi^2$ = Chi square, p = p value*

## Discussion

This study found that about one in 20 of the study participants (5.8%) had a detectable level of serum β-HCG above the WHO limit (5 IU/L). This finding contrast a study conducted in the USA that found that 1.7% of women had HCG levels above 5 IU/L [12]. The disparity in findings can be attributed to the different populations studied. This study focused on women of black African origin, while the USA study examined women of Caucasian and Hispanic origin. The genetic differences between these populations likely contribute to variations in serum β-HCG levels. This finding is in contrast to another study that reported a higher prevalence (8%) of women with levels above the WHO limits [7]. The difference in sample sizes may contribute to this discrepancy. This study’s findings also exceeded the range of serum β-HCG levels reported in a Norwegian study that found a range of 0.1-0.2 IU/L among 732 women below 45 years old [6]. The low levels in the Norwagian study can be explained by variations in menstrual cycle phases, as they examined women during the secretory phase, whereas this study did not consider the menstrual cycle.

This study about one in 25 of the study participants (4.0%) were had suspicious ovarian lesions. The prevalence of ovarian lesions in this study was higher than the reports by a study in the USA that reported a prevalence of 0.04% among 20-39 year old women [16]. The dissimilarity in findings can be attributed to variations in sample size. This study utilized a smaller sample of 224 participants compared to the larger census samples used in the USA study. Furthermore, two other studies in the USA reported higher prevalence rates of 9.2% and 5% respectively [17, 18]. The dissimilarity in findings could be due to the retrospective designs and census sampling methods differ from the cross-sectional and quota sampling approach used in this study, potentially introducing selection bias.

This study found that participants who had elevated serum HCG (> 5IU/L) where 59 folds more likely to have a suspicious ovarian lesion. This finding corroborates a Germany that found significantly higher serum beta-HCG concentrations among women with malignant ovarian tumours compared to those with benign tumours [19]. The similarity in findings was expected since collected data were analyzed at a 5% level of significance and 95% confidence interval

This study found a significant association between serum β-HCG level and age as well as parity. This finding suggests that age is marginally associated with serum HCG concentration. Thus if an individual is older, the possibility of having an ovarian lesion is perhaps more. This finding corroborates with a study that noted that Age was significantly associated with serum HCG levels [7]. Given that both studies were non-experimental studies the similarity in findings were quite expected. This finding corroborates the findings on a pivotal study that found a significant association between parity and serum HCG such that maternal serum β-HCG deceased with increasing parity (p = <0.050).among 20,009 women aged 18-40 years. This finding implies that increasing parity reduces serum HCG and hence protective to the ovaries.

The key limitations of this study are as follows. This study utilized a relatively small quota sample of 224 women, which may not fully represent the larger population of women of reproductive age in Port Harcourt or in Nigeria. The findings may not be generalizable to a broader population, limiting the external validity of the study. The study employed a quota sampling method, which introduces the potential for selection bias. Quota sampling may not provide an equal chance of selection to all members of the target population, leading to a non-representative sample but it was the most feasible method.

## Conclusion

The findings of this study highlight the significance of serum β-HCG levels in detecting suspicious ovarian lesions among non-pregnant women. Approximately 5.8% of the participants had detectable levels of serum β-HCG above the World Health Organization reference, indicating a potential risk for ovarian anomalies. Participants with elevated β-HCG levels were found to be 59 times more likely to have suspicious ovarian lesions compared to those with normal levels. The association between serum β-HCG levels and age as well as parity further emphasizes the importance of monitoring these levels in different demographic groups. Based on these results, it is recommended that non-pregnant women undergo serum β-HCG testing at least once a year to facilitate early detection and intervention for ovarian abnormalities.

## Data Availability

All relevant data are within the manuscript and its Supporting Information files.

## Acknowledgements

We thank the administration of African Centre of Public Health and Toxicological Research (ACE-PUTOR), University of Port Harcourt, Nigeria, for the organizational and administrative support we received throughout this study.

